# Identification of age-associated transcriptomic changes linked to immunotherapy response in primary melanoma

**DOI:** 10.1101/2022.01.23.22269554

**Authors:** Ahmed Ezat El Zowalaty

## Abstract

Melanoma is a lethal form of skin cancers that develops due to constitutive activation of MAPK signaling pathway driven by BRAF and NRAS mutations. Immunotherapeutic agents such as anti-PD-1 (pembrolizumab and nivolumab) and anti-CTLA-4 (ipilimumab) have revolutionized melanoma treatment, however drug resistance is rapidly acquired. Several studies reported the increase in melanoma rates in older patients. Thus, the impact of ageing on transcriptional profiles of melanoma and response to immunotherapy is essential to understand. In this study, bioinformatic analysis of RNA seq data of old and young melanoma patients receiving immunotherapy identified significant upregulation of extracellular matrix and cellular adhesion genes in young cohorts while genes involved in cell proliferation, inflammation, non-canonical Wnt signaling and tyrosine kinase receptor ROR2 were significantly upregulated in the old cohort. Several Treg signature genes as well as transcription factors that are associated with dysfunctional T cell tumor infiltration, were differentially expressed. Differential expression of several genes involved in oxidative phosphorylation, glycolysis and glutamine metabolism has been observed as well. Taken together, this study provides novel findings on the impact of ageing on transcriptional changes during melanoma and novel therapeutic targets for future studies.

## Introduction

Malignant melanoma is a lethal form of skin cancers accounting for more than 9000 deaths per year in the US [1]. Despite the implementation of health measures such as application of sunscreens, the incidence of melanoma is increasing to more than 80000 cases per year in the US [2]. Melanoma development is driven by mitogen-activated protein kinase (MAPK) pathway activation which is driven by mutations in BRAF (52 %) or NRAS (28 %) leading to malignant transformation [3]. The most common BRAF mutations are V600E followed by V600K and V600R [3]. Resistance to BRAF^V600E^ inhibitors after treatment remains a challenge to melanoma treatment. At later stages, melanoma resection remains the main treatment with poor prognosis [4]. The development of immunotherapy has revolutionized melanoma treatment, however, melanoma acquires resistance making treatment more challenging [5]. Aging is an inevitable biological process and a risk factor for many diseases including melanoma. Aging is associated with accumulation of damaged molecules leading to replicative senescence in dividing cells, telomere shortening and reduced or absence of telomerase (hTERT) expression [6]. Senescent cells constantly produce proinflammatory cytokines, chemokines, growth factors leading to a microenvironment favorable for development of age-related diseases including cancer. Several studies have shown that the incidence of melanoma increases with age. Melanoma rates increased by an annual percentage change of 1.8% in adults aged 40 and higher [7]. In addition, there has been a statistically significant decrease in melanoma rates in young adults [7]. Patients aged 65 and older are the most vulnerable age group affected by melanoma-related deaths [8]. Animal experiments have shown that melanoma cell line Yumm1.7 derived from Braf^V600E^/Cdkn2a^−/−^/Pten^−/−^ mouse model of melanoma grows slowly but aggressively in aged mice with increased angiogenesis and metastases due to the age-related increase in the Wnt antagonist sFRP2 [9]. Luckily, old melanoma patents have been found to have better response to the immune checkpoint inhibitor anti-PD1 due to accumulation of a FOXP3^+^ Treg cell population [10]. It has been also observed that in young mice as well as young adults with melanoma, there was accumulation of a significantly higher population of Treg cells driving resistance to immune check point inhibitors [10]. It is therefore important to comprehensively understand the molecular mechanisms driving melanoma progression during aging to identify new therapeutic targets for treatment of malignant melanoma. Elucidating the transcriptional profiles of aging and their association with physiological and pathological changes in melanoma is therefore critical to identify new therapeutic targets. High-throughput genome-wide transcriptomic profiling is a powerful tool that identifies gene expression signatures. The Cancer Genome Atlas (TCGA) is a comprehensive project cataloguing genomic and transcriptomic data of cancers by the National Cancer Institute (NCI), the National Human Genome Research Institute (NHGRI) and the National Institute of Health (NIH) [11]. Since transcriptomic states of aged and young melanoma patients are not fully understood leading to a knowledge gap, in this study, robust bioinformatic analysis of publicly available RNA seq data from 13 patients receiving immunotherapy representative of two distinct age groups, was performed. Five old and eight young patients diagnosed with melanoma were collected and analysis of gene expression profiles was performed. Transcriptomic changes and gene expression signatures associated with response to immunotherapy were investigated.

## Methods

### Clinical cohorts

Two cohorts of old (n= 5) and young (n=8) immunotherapy-treated patients (ipilimumab) diagnosed with primary malignant melanoma were identified from TCGA and served as datasets for evaluation of age-associated transcriptomic changes (Case IDs: supplementary file 1). The class of phenotypes that was used was primary tumor in the clinical category of “primary tumor”.

### Data selection and processing

Transcriptomic RNAseq gene expression level 3 data containing Reads per Kilobase per Million mapped reads (RPKM) were downloaded for each case. RPKM is a widely used RNAseq normalization method and is computed as follows: RPKM= 10^9^(C/NL), where C is the number of reads mapped to the gene, N is the total number of reads mapped to all genes, and L is the length of the gene. RPKM results are generated with SeqWare pipeline. Reference genome is genome build GRCh38.p0, genome name GRCh38.d1.vd1. Alignment of raw reads was done with BWA.

### Differential expression analysis of individual genes

Differentially expressed genes (DEGs) were analyzed using the DESeq2 Bioconductor R package [12]. Normalization was based on the Relative Log Expression method in DESeq2. Identification of significant DEGs was based on a log2 fold change cutoff value of ≤2 (for down-regulated genes) or ≥2 (for up-regulated genes). A significance level of adjusted *P* value of <0.05, using a false discovery rate (FDR) cutoff of <0.1 was set. Heatmaps were generated using “pheatmap”, “RColorBrewer”, “ComplexHeatmap” and “circlize” R packages [13]. Volcano plot was visualized using the EnhancedVolcano R package [14].

### Functional and Pathway Enrichment Analysis

Entrez-ID of each DEG was obtained with the “org.Hs.eg.db” R package then Gene Ontology (GO) and Kyoto Encyclopedia of Genes and Genomes (KEGG) pathway analyses were performed using the “clusterProfiler,” “enrichplot,” and “ggplot2” R packages. KEGG analysis is used to discover pathways enriched in genes in a gene set of interest. Gene set enrichment analysis (GSEA), which is not restricted by DEGs, was also used.

### Gene Ontology and Kyoto Encyclopedia of Genes and Genomes (KEGG) pathway mapping analysis

The Database for Annotation, Visualization and Integrated Discovery (DAVID) was used to analyze the differentially expressed genes. Pathways were also analyzed by the KEGG program. In KEGG pathway enrichment analysis, enriched pathways were identified according to *P* < 0.05.

### Principal component analysis

Principal component analysis was performed with the R packages FactoMineR and Factoextra. Two-dimensional PCA plots were generated using the R ggplot2.

### Statistical analyses

A significance level of adjusted *P* value was set at < 0.05 and a false discovery rate (FDR) cutoff of <0.1 was set. All analyses were performed with R version 4.1.1 (2021-08-10) using the following packages gplots, RColorBrewer, org.Hs.eg.db, statmod, DESeq2, EnhancedVolcano, genefilter, pheatmap, NMF, ggplot2, scales, viridis, gridExtra, reshape2, ggdendro, IHW and dendextend. All graphs were generated in R version 4.1.0.

## Results

### Demographic and clinical features of the patients

Patients with primary diagnosis malignant melanoma (disease type Nevi and Melanomas) were classified into two groups according to age. The young age group had an average age at diagnosis of 42.8±7.75 years. The old group had an average age at diagnosis of 70±4.5 years, *P* = 0.00001 (Figure 1 A). None of the patients displayed prior or synchronous malignancies. Survival analysis showed that the old age group had significantly worse melanoma-specific survival *P* = 0.006 (Figure 1B).

**Figure 1.**
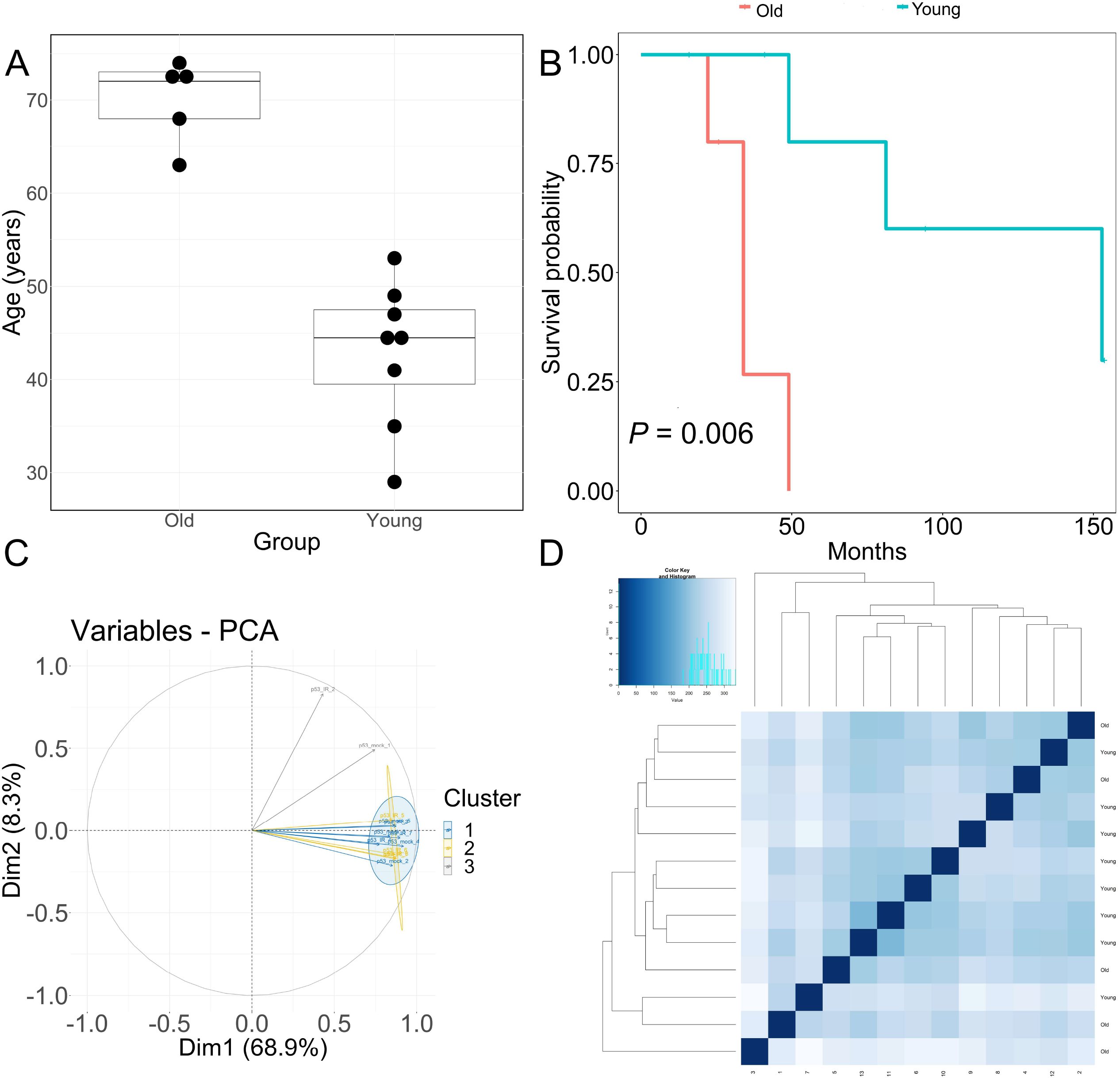
Clinical features of patients and principal component analysis of RNA-seq data. (A) Boxplot of age (in years) of the old and young TCGA patient cohorts used for transcriptomic analysis. *Horizontal line in box* Median, *top, bottom of box* first, third quartile, respectively. (B) Melanoma-specific survival analysis of old and young patient cohorts. (C) PCA analysis plots of RNA-seq samples. (D) Euclidean sample distances heatmap of RNA-seq samples.

### Identification of age-associated differentially expressed genes in melanoma

Quality of RNA-seq data was evaluated with principal component analysis (PCA) and clustering of RNA-seq samples using Euclidean distance. PCA analysis plots of the 13 RNA-seq samples showed clear separation of samples along principal component 1 (PC1) which explained around 70 % of total variance (Figures 1C). In addition, Euclidean sample distances showed that the two age groups are well-clustered (Figure 1D). Differential expression analysis was performed using DESeq2 R package to identify upregulated and downregulated genes between old and young patients diagnosed with primary malignant melanoma. DEGs were visualized as an MA plot (log2 fold change vs. mean of normalized counts) of young vs. old. Red dots represent transcripts with positive and negative log2fold change values also indicating upregulation and downregulation of DEGs (Figure 2A). A total of 3112 significantly DEG genes were identified using a statistical cutoff: q < 0.05. 1345 genes were upregulated, and 1767 genes were downregulated in the young cohort (Figures 2A and 2B). Analysis of DEGs using hierarchical clustering heatmap revealed distinct gene expression profiles between the two age groups (Figure 3A). At cutoff log2 fold change values of 3 and −3, there were 200 upregulated genes and 386 downregulated genes in the young cohort compared to the old cohort respectively. The genes with the lowest *P* values were ATPase Family AAA Domain Containing 3B (*ATAD3B*) and Immunoglobulin Like and Fibronectin Type III Domain Containing 1 (*IGFN1*). Microsomal glutathione S-transferase 1 (*MGST1*), a gene that plays a role in inflammation was remarkably upregulated in the young age group (log2 fold change = 3.02) (Figure 3B). Four collagen genes involved in extracellular matrix and cellular adhesion were upregulated in the young group, COL14A1, COL2A1, COL6A6, COL4A4 (Figure 3B). We also observed significant upregulation of cell proliferation genes p21^CIP1^ (CDKN1A) and CDK2 in the young cohort, while CDK1 was upregulated in the old cohort *P* < 0.005 (Figure 3C). Slight upregulation of ARMC8 was also observed in the young cohort (Figure 3C). *ARMC8* plays critical roles in cell proliferation, apoptosis, differentiation and has been reported to be a prognostic marker in several malignancies including liver, lung and breast cancers [15-18]. It has been reported that *ARMC8* is upregulated in malignant melanoma cell lines and was associated with increased invasiveness of melanoma [15]. We also observed significant upregulation of Interleukin-17A (*IL*-*17A*) and interleukin 11 (IL11) in the old cohort *P* < 0.05 (Figure 3D). Interleukin-17A (*IL*-*17A*) is produced by Th17 cells which infiltrate the tumor microenvironment and induces expression of several inflammatory cytokines including IL-1β, IL-16 and IL-23 leading to variable effects on tumor growth at different stages [19, 20]. IL-17A upregulation has been reported in a 53-year-old male with a collision primary laryngeal malignant melanoma and invasive squamous cell carcinoma [21]. IL-11 is a pleiotropic interleukin involved in tumor development. IL-11 has been shown to be elevated in colorectal cancer and in the exosomes of metastatic uveal melanoma and associates with poor prognosis in melanoma [22-25]. It is known that phenotype switching is characteristics of melanoma, where proliferative cells tend to be non-invasive, while invasive cells tend to be non-proliferative, a process regulated by Wnt signaling pathway [26-28]. We identified the dysregulation of several genes in the Wnt signaling pathway (Figure 3E). Interestingly, the non-canonical Wnt molecule Wnt5A and the tyrosine kinase receptor ROR2 which drive invasion, metastasis and therapeutic resistance [29-31], were significantly upregulated in the old cohort (Figure 3E).

**Figure 2.**
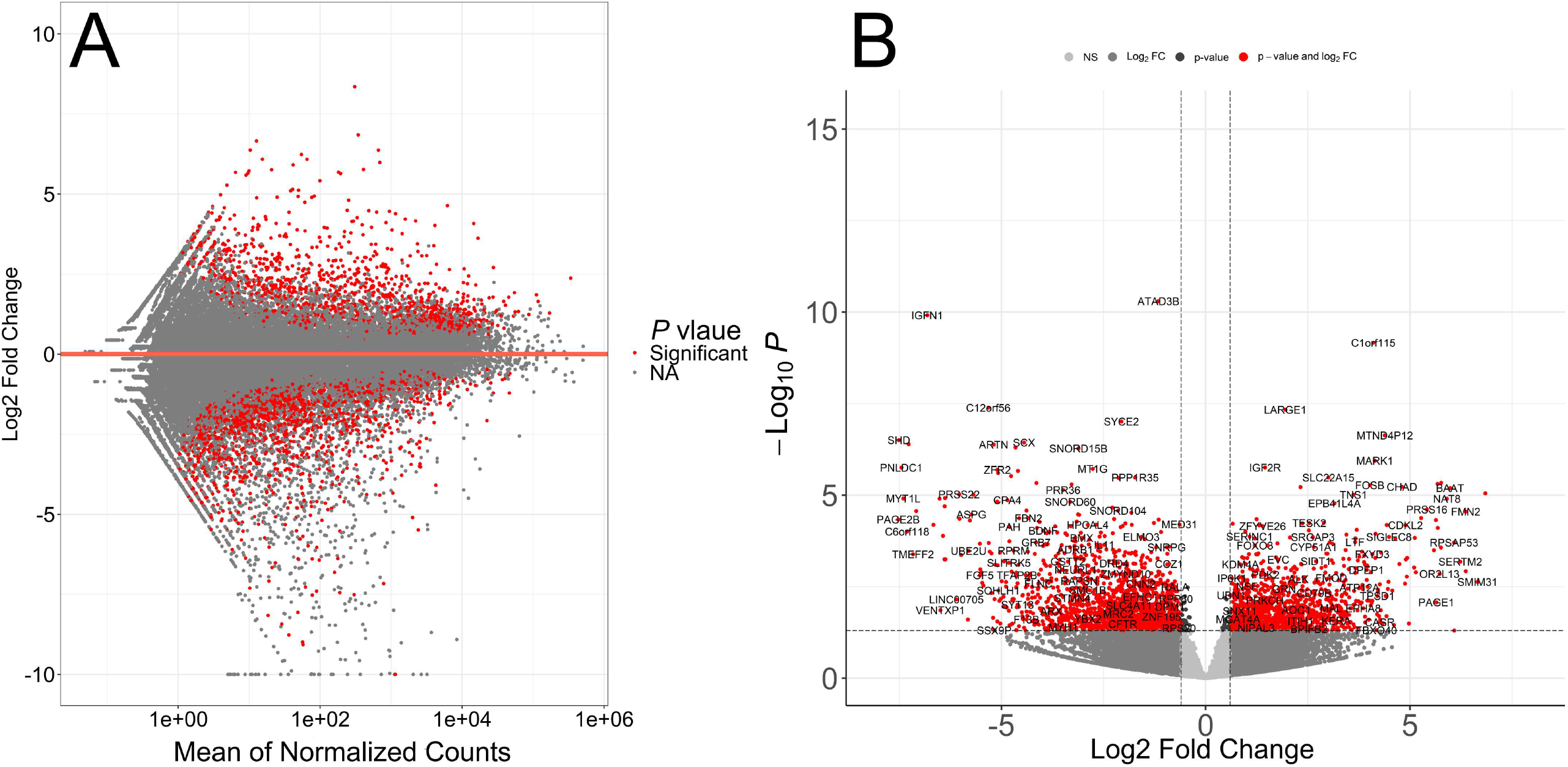
Transcriptomic analysis of TCGA patient’s RNA-seq data. (A) MA plot displaying the log2 fold-change compared with mean of normalized counts generated with DESeq2 package. (B) Volcano plot of differentially expressed genes. Direction of comparison Young/Old, 1345 genes were upregulated and 1767 genes were downregulated. Red: Significant, Grey: Not significant. Right: Upregulated, Left: Downregulated.

**Figure 3.**
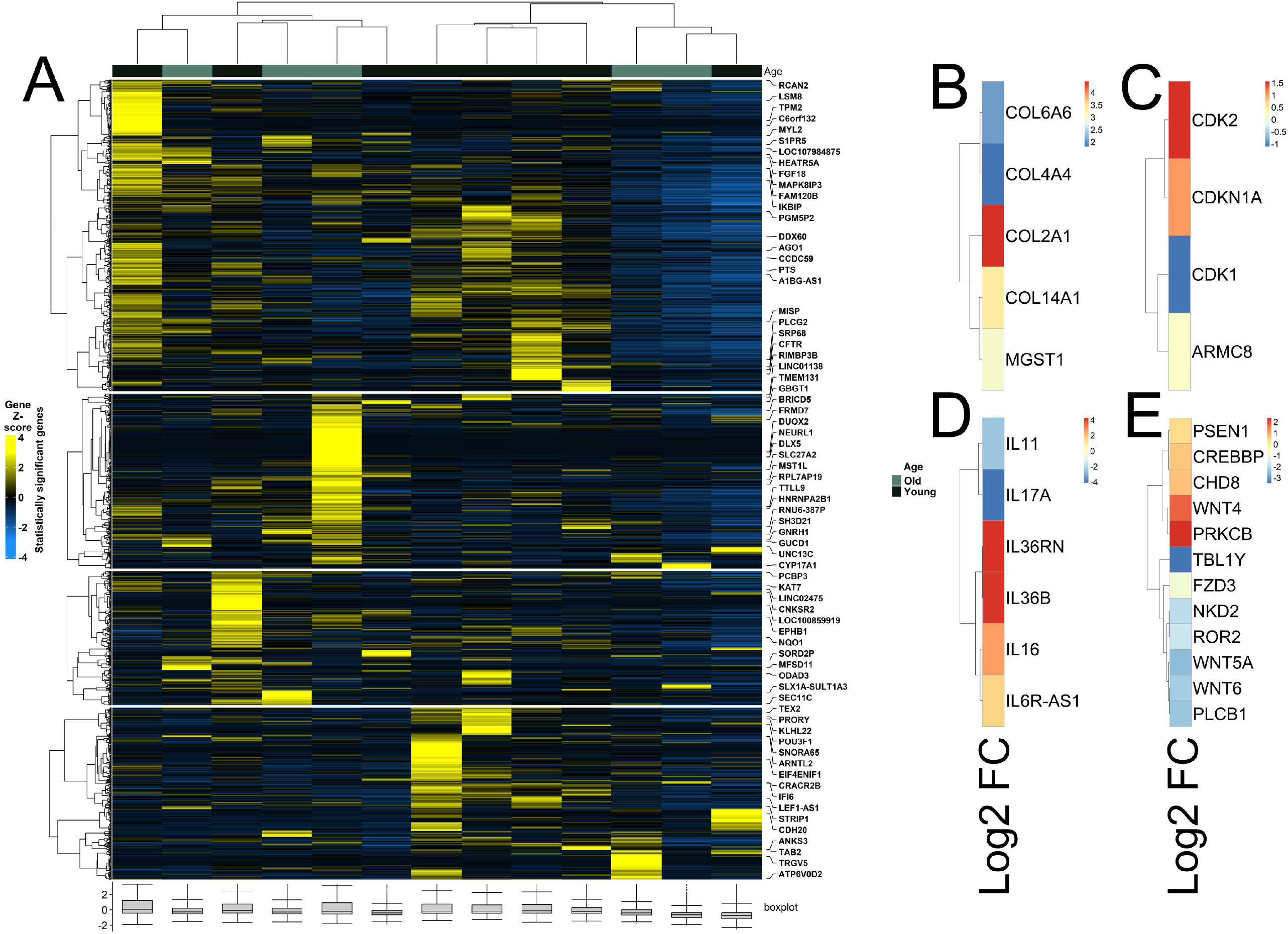
Heatmaps of differently expressed genes between young and old patient cohorts. (A) Hierarchical clustering heatmap of Gene Z score showing significantly differentially expressed genes between young and old age groups. (B) Differential expression of *MGST1* and collagen genes. (C) Differential expression of *cell proliferation* genes. (D) Differential expression of *interleukin* genes. (E) Differential expression of genes of Wnt signaling pathway.

### Pathway analysis, functional annotation of differentially expressed genes and identification of key regulatory genes of response to immunotherapy

Functional annotations clustering of the most upregulated and downregulated genes (log2 fold change =3 and −3 respectively) was performed with DAVID to identify the biological processes that are related to differential gene expression changes in the two melanoma age groups receiving immunotherapy. Among the 200 genes upregulated in the young group, the top clusters were related to signal peptide, Glycoprotein, glycosylation site:N-linked (GlcNAc) and Ion transport. The top clusters in the old age group were related to extracellular region, signal peptide, extracellular space and glycosylation site:N-linked (GlcNAc). Pathway and gene set enrichment analysis of the upregulated genes showed significant enrichment of genes involved in several biological processes most notably interferon-α, interferon-γ response, IL2-STAT5 signaling, JAK-STAT3, P53 and inflammatory response pathways (Figure 4A). KEGG pathway analysis of DEG genes was also performed and revealed that pathways that had the highest enrichment scores for the DEG genes were riboflavin metabolism (0.71), Histidine metabolism (0.64), Antigen processing and presentation (0.6), Toll-like receptor signaling, (0.6), Circadian rhythm (0.57), Lysosome (0.55), calcium signaling and PD-L1 expression/PD-1 checkpoint pathway in cancer (Figures 4B and 4C). It is known that cancer cells upregulate programmed death ligand-1 (PD-L1) that binds inhibitory receptor programmed death receptor-1 (PD-1) on T cell surface to avoid immune attack [32]. Several studies have evaluated the efficacy of PD-1 and PD-L1 inhibitors in different cancer including lung cancer, renal cancer and malignant melanoma in PD-L1–negative and PD-L1–positive tumors. Therapeutic benefit from immunotherapy was observed in patients within the PD-L1–negative tumors, however, resistance to immunotherapy remains a challenge [33, 34]. Indeed, gene set enrichment analysis (GSEA) of transcriptomic data confirmed the significant enrichment of genes in the T cell receptor signaling, PD-L1 expression and PD-1 checkpoint and JAK-STAT signaling pathways which are essential pathways in melanoma (Figures 4D-4G) [35-37].

**Figure 4.**
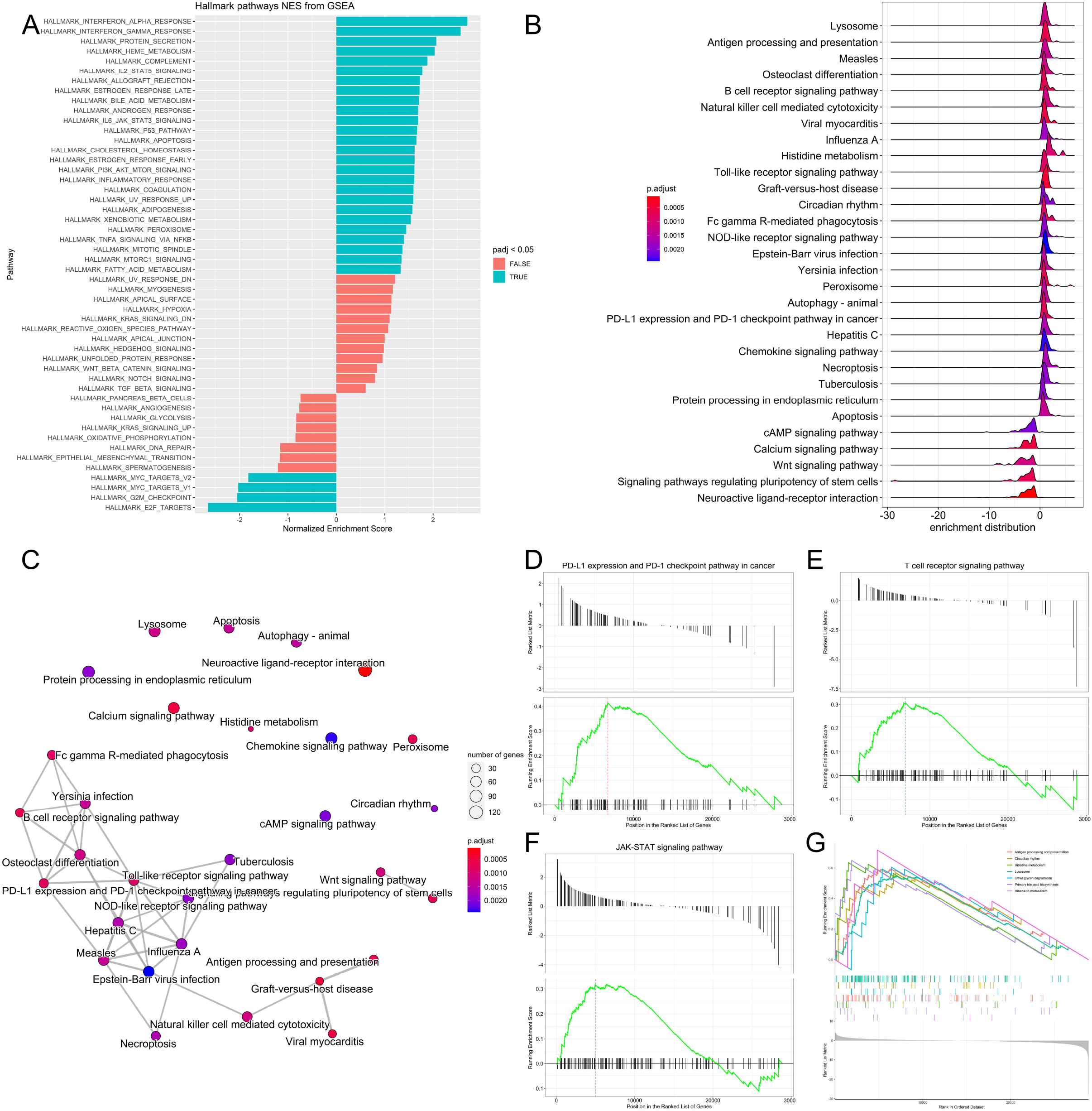
Pathway analysis, functional annotation of differentially expressed genes and identification of key regulatory genes of response to immunotherapy. (A) Barplot showing biological process terms and pathways of GSEA. GSEA normalized enrichment scores are presented on the *x*-axis. (B) GSEA ridge plot for of differentially expressed genes, the gradient indicates low to high adjusted *P* values. (C) Functional enrichment analysis from GSEA results. Nodes represent each enriched gene set of canonical pathways. Downregulated molecules shown in shades of blue and upregulated genes are shown in shades of red. (D-G) Enrichment plots of top regulatory gene networks identified using gene set enrichment analysis. Significance was set at *P*-values (≤0.05) and false discovery rate (≤ 0.25). Enrichment plots show running enrichment score of the gene set as a running-sum statistic working down the ranked list of genes. Red dashed line is the enrichment score peak of the plot for a specific gene set. Black vertical bars are positions of genes in regulatory gene subnetworks in the ranked list of genes. Leading edge is gene subset contributing mostly to enrichment scores and are the most differentially expressed genes.

### Differential expression of Treg signature genes and metabolic genes in melanoma

*FOXP3*^+^ T regulatory cells (Tregs) are characterized by expression of the transcription factor forkhead box P3 (*FOXP3*). They infiltrate the tumor microenvironment thus providing an immunosuppressive microenvironment and immune tolerance [38]. Functionally immunosuppressive Tregs are also highly enriched in the tumor microenvironment of patients with melanoma [39-41]. Tregs provide an immune-tolerant microenvironment by secreting a number of factors such as IL-10, TGF-β, IL-35, PD-1, LAG-3 and TIM-3 [42, 43]. Inhibition of Treg function is therefore required for tumor clearance and therapeutic effectiveness. It has been shown in a cohort of primary melanoma samples from NYU and Vanderbilt, that young patients acquire resistance to immunotherapy due to reduction in *FOXP3*^+^ Treg cell population, while old patients (>66 years) have low number of CD8+ T cell population [10]. Animal experiments in murine models of melanoma showed that depletion of Tregs led to tumor clearance and increased survival indicating the critical importance of this cell type for antitumor immunity [44]. Thus, in this analysis, Treg signature genes in young and old patients treated with immunotherapeutic agent ipilimumab were compared to dissect the molecular pathways regulating T cell function in melanoma during aging. Two Treg-signature genes, *NR4A3* and *IKZF2* were significantly upregulated (log2 fold change > 1.5) (Figure 5A). Nr4a3 is an orphan nuclear receptor upregulated in T cells undergoing differentiation and is required for *FOXP3* induction in T cells [45-48]. *IKZF2* (Helios) is a transcription factor required for suppression of IL-2 production in Tregs and regulates *FOXP3* binding to the *IL2* promoter thus silencing *Il2* transcription in Tregs. [49]. The expression of the most prominent signature genes of tumor infiltrating Tregs such as *CXCR5, IL17F, IL17A, IL22, FOXP3, IL12RB2, TNFRSF9, CD274, TNFRSF4, TNFRSF9, IL10, CCR7* and *STAT1* was also investigated (Figure 5A). ID3, RDH10 and EOMES are transcription factors associated with a dysfunctional tumor infiltrating T cell state [50]. ID3 and RDH10 were enriched in the old cohort while *EOMES* was enriched in the young cohort (Figure 5A). We also observed significant upregulation of *STAT1* in the young cohort (Figure 5A). *STAT1* is involved in cytokine production and is required for recruitment and activation of T cells in tumor microenvironment. STAT1 has also been found through a 2 cell type-CRISPR screen including human T cells as effectors and melanoma cells as targets, to be upregulated affecting immunotherapy response [51]. The GTP-binding protein 4 (*GBP4*) was previously identified as a prognostic signature gene that separates melanoma patients into low and high risk groups according to survival [52]. *GBP4* was significantly upregulated in the young group (Figure 5A). In addition, we examined metabolic gene expression in young and aged cohorts. It is well known that metabolic rewiring is a hallmark of all types of cancers [53-55]. It has been previously shown that immunotherapy (PD-1 blockade)-resistant melanoma tumor cells acquire hypermetabolic state by upregulation of glycolysis and mitochondrial oxidative phosphorylation. Upregulation of lactate and TCA cycle metabolites is observed in immunotherapy-resistant melanoma cells [56]. Glutamine addiction is a hallmark of malignant transformation including melanoma [57]. Glutamine is an anaplerotic amino acid. Carbon derived from glutamine is used to maintain TCA cycle intermediates and its nitrogen is used for transamination reactions, purine and redox intermediates NAD and NADP biosynthesis [58]. Upregulation of genes involved in biosynthesis of proline from glutamate is also a characteristic of malignant melanoma [59, 60] and depletion of glutamine sensitizes melanoma cells to TRAIL-mediated cell death [61]. Five genes *ATP12A, ATP6AP1, ATP6V0A1, ATP6V0C and ATP6V0D2* involved in oxidative phosphorylation pathway were upregulated in the young cohort (Figure 5B) while six genes *ATP6V1C2, COX6C, COX7B, PPA1, UQCRB and UQCRH* were upregulated in the aged cohort (Figure 5B). Glycolysis genes *ALDH9A1, LDHAL6A, PFKL* and *PKLR* were upregulated in the young cohort (Figure 5B), while the glutamine metabolism gene *GLS2* was notably upregulated in the aged cohort (Figure 5B). Dysregulation of (*NFE2L2* or *NRF2*) target genes was also observed. The transcription factor Nuclear Factor Erythroid 2-Related Factor 2 (*NFE2L2* or *NRF2*) is the master regulator of cellular redox homeostasis. *NRF2* regulates the expression of genes involved in antioxidant defense and xenobiotic metabolism. *NRF2* is activated in melanoma, required for melanoma cell proliferation and its knockdown sensitized melanoma cells to oxidative stress [62]. *NRF2* expression was slightly upregulated in the young cohort while its target genes *NQO1* and *ALAS1* were significantly upregulated in the young cohort. *GSTT2B, GSTT2* and *TXNDC17* were significantly upregulated in the old cohort (Figure 5B). Finally, analysis of correlation between gene expression and melanoma patients’ survival from TCGA showed that *CCR7* (Hazard Ratio (HR) 0.67, *P* = 0.0029), *CXCR5* (HR 0.7, *P* = 0.01), *EOMES* (HR 0.76, *P* = 0.047), *GBP4* (HR 0.5, *P* = 4.9e-07), *TNFRSF9* (HR 0.59, *P* = 0.00011) and *STAT1* (HR 0.58, *P* = 5.3e-05) were significantly related to patient prognosis. Overall survival rate of patients with high expression of *CCR7, CXCR5, EOMES, GBP4, TNFRSF9* and *STAT1* in melanoma was significantly higher (Figures 5C – 5H) which is consistent with the significantly higher survival rate observed for the young cohort receiving immunotherapy (Figure 1B).

**Figure 5.**
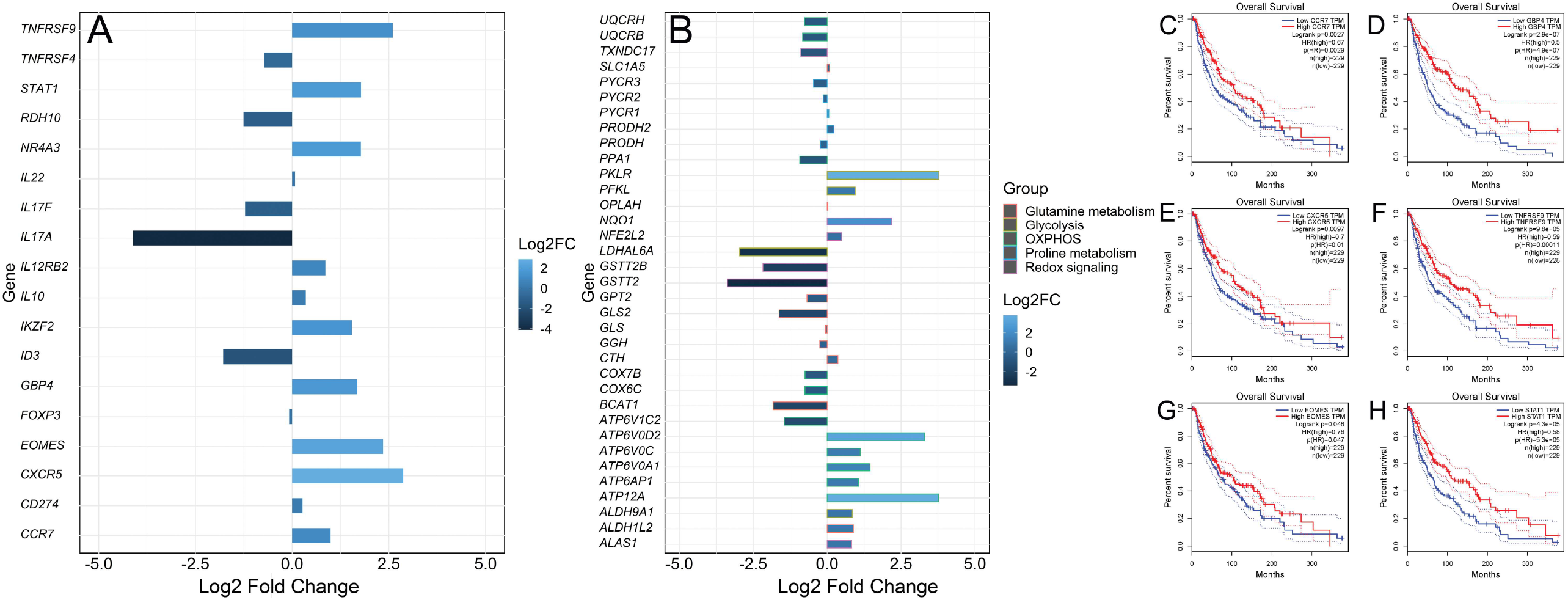
Differential expression of Treg and metabolic genes and correlation analysis of gene expression and patients’ survival from TCGA. (A) Bar plot of upregulated and downregulated Treg signature genes. (B) Bar plot of upregulated and downregulated metabolic genes. (C-H) Survival curves of TCGA patients that express low and high levels of *CCR7, CXCR5, EOMES, GBP4, TNFRSF9* and *STAT1*.

## Discussion

Melanoma accounts for 80% of skin cancer deaths [63]. Although, immunotherapy has revolutionized melanoma treatment, many melanoma patients remain resistant to immunotherapy [64, 65]. Immune checkpoint inhibitor proteins such as CTLA-4 and PD-1 are expressed in activated T cells. CTLA-4 inhibits activation of T cells and PD-1 binds to PD-L1/PD-L2 ligands that are expressed in melanoma, thus inhibiting immune attack [63]. Aging is a risk factor for melanoma incidence. In this study, the impact of aging on the transcriptomic profiles of melanoma patients receiving immunotherapy was investigated. Comprehensive analysis including gene expression, identification of DE genes, GSEA and pathway enrichment was performed to identify signature genes. Our results show that the old age cohort had a significantly worse melanoma-specific survival *P* < 0.001 (Figure 1A). 1345 DE genes were upregulated, and 1767 DE genes were downregulated in the young cohort (Figures 2A and 2B). The results reveal signatures of the impact of age on melanoma. *MGST1* which plays a critical role in inflammation was remarkably upregulated in the young cohort. Collagen genes COL14A1, COL2A1, COL6A6, COL4A4 involved in extracellular matrix organization were upregulated in the young group (Figure 3B). Genes regulating cell proliferation, apoptosis and differentiation were upregulated in the young cohort (Figure 3C). Activation of inflammatory genes such as *IL*-*17A, IL-11* was observed in the old cohort (Figure 3D). *IL*-*17A* induces the expression of inflammatory cytokines IL-1β, IL-16 and IL-23 in the tumor microenvironment in malignant melanoma [19-21]. Activation of such inflammatory pathways is potentially associated with the poor survival observed for the old cohort receiving immunotherapy. It has also been reported that phenotype switching in melanoma is regulated by the Wnt signaling pathway and that Wnt5A-treated B16 melanoma cells acquire greater metastatic and invasive potential, an effect mediated by the orphan tyrosine kinase receptor ROR2 [30, 66]. Indeed, Wnt5A and ROR2 were upregulated in the old cohort receiving immunotherapy (Figure 3E). Our results further reveal enrichment of genes in several biological processes including interferon-α, IL2-STAT5 signaling, JAK-STAT3 and P53 pathways (Figure 4A). Interestingly, KEGG pathway analysis of DE genes showed upregulation of PD-L1 expression/PD-1 checkpoint pathway in cancer (Figures 4B-4D). DE gene analysis also revealed differential expression of several Treg signature genes and metabolic genes. Immunosuppressive FOXP3^+^ Tregs infiltrate the tumor microenvironment inhibiting immune attack affecting patient response to immunotherapy [10, 67, 68]. Treg signature genes *CXCR5, TNFRSF9, CCR7, STAT1, GBP4*, and *EOMES* were significantly upregulated in the young cohort and correlated with higher survival in melanoma (Figure 5B and 5C-5H) while *ID3, IL-17A, IL-17F, RDH10* and *IL-11* were significantly upregulated in the old cohort (Figure 5A). We also observed differential expression of several genes involved in metabolic rewiring in melanoma. Oxidative phosphorylation genes *ATP12A, ATP6AP1, ATP6V0A1, ATP6V0C and ATP6V0D2* were upregulated in the young cohort (Figure 5B) while *ATP6V1C2, COX6C, COX7B, PPA1, UQCRB and UQCRH* were upregulated in the aged cohort (Figure 5B). *GLS2* was upregulated in the aged cohort. It has been reported that *GLS2* upregulation increases tumor metastasis and correlates with poor survival [69]. In conclusion, our data reveal transcriptional changes associated with age, dysregulation of many biological, inflammatory and metabolic pathways associated with greater metastatic and invasive potential in old melanoma patients receiving immunotherapy as well as potential novel therapeutic targets in melanoma. One limitation in the current study is the small sample size due to limited data on melanoma patients receiving immunotherapy in the TCGA database. In the future, large samples, clinical cohorts and basic experiments will be required to verify the impact of ageing on immunotherapy response and prognosis of melanoma patients.

## Data Availability

All data produced in the present study are available upon reasonable request to the authors

## Author Contributions

A.E.Z. conceived, designed the research, analyzed the data and wrote the manuscript.

## Ethical approval

Not applicable.

## Data Availability

The datasets of the current study analyzed with R are available from the corresponding author on request.

